# The Mediating Role of SES on the Relationship between Pregnancy History and Later Life Cognition

**DOI:** 10.1101/2021.09.29.21264303

**Authors:** Averi J. Giudicessi, Ursula G. Saelzler, Aladdin H. Shadyab, Alexander Ivan B. Posis, Erin Sundermann, Sarah J. Banks, Matthew S. Panizzon

## Abstract

**Objective:** The association of pregnancy with later life cognition is not well understood. Few studies address the potential confounding role of socioeconomic factors on this relationship. We examined whether pregnancy was associated with cognitive function in a large, population-based sample of post-menopausal women and the potential mediating effects of education level and federal income-to-poverty ratio (PIR) on this relationship.

**Methods:** Participants were 1,016 post-menopausal women from the National Health and Nutrition Examination Survey (NHANES). We utilized data from two study waves between years 2011-2014. Cognitive functioning was evaluated by: Digit Symbol Substitution Test (DSST), Animal Fluency (AF), Consortium to Establish a Registry for Alzheimer’s Disease CERAD word learning task (CERAD-WL) and CERAD delayed recall (CERAD-DR). Lifetime education level and federal income-to-poverty ratio (PIR) were examined as mediating factors. Regression models were used to examine the relationship between number of term pregnancies and incomplete pregnancies and cognitive performance.

**Results:** A greater number of term pregnancies was related to worse performance on the DSST (p < .001), CERAD-DR (p < .007), and AF (p < .03). Conversely, greater incomplete pregnancies related to better CERAD-DR performance (p < .03). Significant associations between term pregnancies and cognitive scores were mediated by PIR but not education level.

**Conclusions:** Higher number of term pregnancies was associated with worse cognitive performance, whereas higher number of incomplete pregnancies was associated with better cognitive performance. Results indicate the necessity to consider SES factors when studying the relationship between pregnancy and cognition.

## INTRODUCTION

Women have higher rates of Alzheimer’s disease and related dementias (ADRD) than men^1^. Almost two-thirds of Americans living with AD are women.^1,2^ Reproductive events that are unique to women (e.g. menarche, childbearing, menopause) and their relation to later life cognition have gained increasing attention in recent literature and might contribute to sex differences in ADRD risk.^3–8^ Pregnancy results in a myriad of physiological changes that are known to impact brain health including fluctuation in hormone levels, neuroplasticity, immune function, and cardiac output.^9–12^ However, the association between pregnancy and later life cognition is poorly understood.

The majority of research on reproductive history and late-life cognition to date has taken an estrogen focused approach that assumes that longer estradiol exposure is beneficial to cognitive function. This would suggest a positive relationship between pregnancy history and cognitive function as estrogen levels rise during pregnancy. However, the relationship between pregnancy history and cognition has been inconsistent.^3–5,13^ For example, in a longitudinal study of 361 postmenopausal women, those who reported having never given birth to a live child exhibited better global cognitive functioning over 12 years than women who reported having given birth to a live child.^13^ Similar studies suggest that grand multiparity, defined as having 5 or more children, has a negative effect on cognition as measured by Mini-Mental State Examination (MMSE), Digit Symbol Substitution Test (DSST), and DSM-IV criteria for dementia diagnosis.^4,5,7,14^ Alternatively, Lange et al^3^ found that women with prior pregnancies had slower brain aging in striatal and limbic regions, with strongest associations in women with 5 or more pregnancies.^3^ Other studies have similarly found better cognitive function or no difference in cognition when comparing women that report term pregnancies with those that report never being pregnant.^15–17^

In addition to term pregnancies, research suggests that the number of incomplete pregnancies may impact later-life cognition.^5,18^ Exposure to estrogen will be different for a woman who has had term pregnancies versus incomplete pregnancies. During pregnancy, estrogen steadily increases in women and reaches its peak level in the third trimester.^19,20^ After a woman gives birth estrogen drops and eventually (roughly 6 months after childbirth) returns to pre-pregnancy levels.^19,21^ Pregnancy may also influence brain and cognitive health via changes to inflammatory response in the body. For example, alterations in the production of T regulatory cells (T_reg_) (cells that play a role in regulating inflammation) in pregnant women varies by pregnancy trimester. T_reg_ cells peak in the first two trimesters and stay stable across late pregnancy and slightly increase postpartum.^22,23^ Taking into consideration these changes at specific points during a pregnancy, it is important to understand if a pregnancy was term or incomplete. This is especially crucial when looking at number of pregnancies within the context of cognitive function. To our knowledge only two previous studies have explored the differences between incomplete and term pregnancies on later-life cognition and both studies found a positive relationship between incomplete pregnancies and cognition as measured by the MMSE and Clinical Dementia Rating (CDR).^5,18^ Jang et al, found that women who had incomplete pregnancies performed better on the MMSE than those women that did not have report an incomplete pregnancy. Furthermore, women who had 5 or more term pregnancies exhibited significantly worse performance on the MMSE than those women that reported between 1-4 term pregnancies.^5^

The role of socioeconomic status (SES) in the association of pregnancy with cognition is an important, yet understudied, research question given SES links to both number of pregnancies and cognition.^24,25^ Early life education has been shown to be associated with cognitive outcomes in later life, and there is a connection between early education and adult SES as more access to education is indicative of access to higher paying jobs and higher income levels in later life.^26^ Lower SES for example: lower income levels, lower education levels, and neighborhood disadvantage, has been associated with a younger age at first pregnancy, higher rates of pregnancy, as well as a higher number of complications during pregnancy that have been linked to poor cognitive outcomes later in life.^27–29^ Women with low SES generally have children at a younger age and experience higher rates of unintended pregnancy.^25,28^ Having a child at a younger age makes access to education even more difficult. Previous research demonstrates a negative relationship between education level and number of children.^29^ Given that education level and other SES markers relate to both number of pregnancies and later life cognition, it is possible that these variables could mediate the relationship between number of pregnancies and cognitive function among women, especially those who had a greater number of children.

The objective of this study was to examine the association of number of full-term and incomplete pregnancies with later life cognition among postmenopausal women from National Health and Nutrition Examination Survey (NHANES). Based on the existing literature, we hypothesized that a greater number of pregnancies would be associated with low cognitive function and a greater number of incomplete pregnancies would be associated with higher cognitive functioning. We also explored the potential mediating effects of education level and federal income-to-poverty ratio (PIR) on this relationship.

## METHODS

### Study Population

Data were obtained from the National Health and Nutrition Examination Survey (NHANES). Started in 1999, the NHANES is a cross-sectional, population-based study that aims to understand the health and nutritional status of a nationally representative sample of adults and children living in the United States. In each wave, around 5,000 participants are enrolled. The survey is administered by the National Center for Health Statistics (NCHS), a part of the Centers for Disease Control and Prevention (CDC). Information is collected through participant interviews and health examination. We utilized data available for two waves collected during 2011-2014. Participants who did not have demographic, reproductive history, or complete neuropsychological data, and women who had 12 or more pregnancies (N=5) were not included in the final analysis; 5,547 women were excluded for this reason. The final analytic sample included 1,016 women.

### Measures

Per the NHANES protocol, participants aged 60 years and older were eligible to complete the cognitive assessment. Cognitive tests were completed in the participants home. Participants who understood or read English, Spanish, Korean, Vietnamese, traditional or simplified Mandarin, or Cantonese were administered measures in their native language. Three cognitive tests were administered to participants: Digit Symbol Substitution Test (DSST), Animal Fluency (AF) and Consortium to Establish a Registry for Alzheimer’s Disease (CERAD) Word Learning Subtest and Delayed Recall Subtest (CERAD-WL/CERAD-DR).

#### DSST

This test requires participants to match symbols to numbers according to a key located at the top of the page. The participant is tasked with writing down the corresponding symbol with each digit as fast as they can in a two-minute time frame. The score is calculated as the total number of correct matches. This DSST assesses sustained attention and processing speed, and is sensitive to cognitive deficits in a wide range of brain diseases and conditions.^30^

#### AF

Participants are asked to name as many animals as possible in one minute. A point is given for each animal named within the one-minute time frame. This exercise has been shown to differentiate between individuals with normal cognitive functioning, those with mild cognitive impairment, as well as more severe forms of cognitive impairment such as AD.^31,32^

#### CERAD-WL/CERAD-DR

This test consists of three consecutive learning trials, and a delayed recall. Following the NHANES administration instructions, for the learning trials, participants were instructed to read aloud 10 unrelated words, one at a time, as they were presented. Immediately following the presentation of the words, participants were asked to recall as many words as possible. The words for the learning trials were presented in large bolded format on a computer monitor. However, participants who were unable to read the words on the computer monitor were read aloud the words by a NHANES interviewer and then asked to repeat each word after it was read. The order in which the words were presented changed in each trial. Approximately 8-10 minutes after the last learning trial, the participants were administered the delayed recall trial in which they were again asked to recall all the words in the list as they could remember. The maximum score for each trial is ten.^33^ A total score for the three learning trials and a total for the delayed recall trial were calculated for our purposes. The test was originally developed to assess patients suspected of having progressive memory problems, and the delayed recall portion in particular has been shown to effectively distinguish between individuals with dementia and their cognitively normal counterparts.^34^

### Pregnancy

To calculate number of pregnancies we utilized the question: how many times have you been pregnant? Number of term pregnancies was calculated by adding the total number of reported vaginal and cesarean deliveries. We calculated number of incomplete pregnancies by taking the difference between the questions: 1) how many times have you been pregnant? And 2) the total number of reported vaginal and cesarean births.

### Covariates

Sociodemographic characteristics included age at screening, race, length of reproductive span that was calculated by the taking the difference between the self-reported ages at first and last period. Body Mass Index (BMI) was calculated by trained technicians affiliated with the NHANES study using collected height and weight measurements the day of data collection. Smoking status was defined by whether or not participants reported having at least 100 cigarettes in one’s life. The Patient Health Questionnaire (PHQ) was administered to determine the frequency of depressive symptoms over the past 2 weeks.^32^ Medical conditions such as diabetes, heart disease, thyroid disease, history of stroke, sleep disorders, and hypertension were determined by self-report measures. A composite medical score was calculated from these self-reported variables as the sum of medical conditions.

Federal income-to-poverty ratio (PIR) (calculated per Department of Health and Human Services guidelines as the proportion of total family income to the poverty level; a smaller family PIR suggests a lower family income level) and education level were also used as covariates in our regression models and mediators in our formal mediation analysis.

### Availability of Data and Materials

Data used in this study and corresponding protocols on data collection is freely available on the National Health and Nutrition Survey website. https://www.cdc.gov/nchs/nhanes/index.htm

### Statistical analysis

Analyses were conducted using the R Statistical language (version 1.4.1106).^35^ Term and incomplete pregnancies were run as both continuous and categorical variables. Consistent with the literature, number of term pregnancies were categorized as 0, 1-2, 3-4, or 5 or more and number of incomplete pregnancies were categorized as 0, 1-2, 3 or more.^4,5,14^ Reference groups for our regression models using term and incomplete pregnancies as categorical variables were: zero term pregnancies and zero incomplete pregnancies. We used ANOVA and chi-square tests to examine differences in study characteristics by categories of term pregnancies. Statistical significance was regarded as two-sided *p* < 0.05.

General regression models using the R package gtsummary were used to determine associations of term and incomplete pregnancies with cognitive function measures.^36^ Each cognitive measure (DSST, AF, CERAD-WL, CERAD-DR) was examined in a separate model. Initial models included term and incomplete pregnancies as our exposure variables. Covariates included: age at screening, race, length of reproductive span, BMI, smoking status, number of medical conditions, and depressive symptoms (Model 1). We conducted preliminary tests of mediation by education level (Model 2) and by PIR (Model 3) by systematically including them in the model. Finally, we added both education level and PIR together in a separate model to understand the potential mediating effect of these two variables together on the pregnancy and cognition (Model 4).

Formal mediation analyses were performed using the Imai et. al^37^ approach that allows for the calculation of causal mediation effects without reference to a specific statistical model and allows for an accommodation of both continuous and discrete mediators.^37^ This is important as our education variable is a categorical variable with the following levels: Less than 9th grade, 9-11th grade, High School/GED, some college, and college graduate or above. This model allows for the calculation of indirect effects, average direct effects, total effect and proportion mediated. In the mediation analysis, we fit regression models using term pregnancies as the exposure variable and each specific cognitive test as the outcome variable. We adjusted the models using the following covariates: incomplete pregnancies, age at screening, race, length of reproductive span, BMI, smoking status, number of medical conditions, and depressive symptoms. The R package mediation was used to estimate these effects.^38^ The 95% CI for the proportion mediated are based on 1000 bootstrap samples.

## RESULTS

This study included 1,016 women whose mean age at the time of cognitive assessments was 67.31 (SD = 5.36). Thirty-two percent of the sample reported some college education. Twenty percent of our sample self-described themselves as Hispanic, 46% as White, 24% Black, 8.9% Asian, and 1.4% Multi-Cultural. Eleven percent (N=116) of participants reported never having a term pregnancy and 89% (N = 900) reported at least 1 term pregnancy. Approximately 37% of our sample had reported experiencing at least one incomplete pregnancy. Significant differences between term pregnancy groups were found in the following variables: age, education level, PIR, BMI, and medical conditions. Some of our strongest differences were found in between term pregnancy groups in our education level and PIR variables. For example, 41% of women that reported having no term pregnancies reported graduating college or above while only 6.4% of women with 5 or more term pregnancies reported having graduated college or above. Women who reported having no term pregnancies or 1-2 term pregnancies, reported an average PIR of 2.78-2.95 (SD = 1.69) while those women who reported having 5 or more term pregnancies reported an average PIR of 1.68 (SD=1.21). A comparison of sample characteristics by term pregnancy category is displayed in Table 1.

### Results for Term Pregnancy

We found a significant relationship between term pregnancies and DSST scores. In Model 1 (adjusted for age at screening, race, length of reproductive span, BMI, smoking status, number of medical conditions, and depressive symptoms), higher number of term pregnancies were associated with 0.09-point lower DSST score, (*p* < .001), however this relationship was attenuated, when adding education and PIR into our model (β = -0.01 to 0.03, *p* = 0.017 to 0.052). Higher number of term pregnancies were also associated with a lower AF score (β = -0.03, *p* = .028); however this association diminished after accounting for education and PIR (β = -0.03 to 0, *p* = 0.2 to 0.9). CERAD-WL scores and the relationship with term pregnancies showed a non-significant relationship across all models. (β = -0.02 to 0.01, *p* = 0.2 to 0.6). Finally, higher number of term pregnancies were associated with lower CERAD-DR scores (β = -0.04, *p* = .007) but as was the case with DSST and AF scores, this relationship was significantly diminished when accounting for education level and PIR (β = -0.03 to -0.01, *p* = 0.049 to 0.4).

### Results for Incomplete Pregnancy

There were no significant associations between incomplete pregnancies and DSST across all models (β = 0 to 0.01, *p* = 0.7 to 0.9). Furthermore, incomplete pregnancies were not associated with significantly different AF scores across models (β = 0.06, *p* = 0.034 to 0.59). The association of incomplete pregnancies with CERAD-WL was not significant across all models (β = 0.05, *p* = 0.085 to 0.11). Finally, incomplete pregnancies were positively associated with CERAD-DR across all models: (β = -0.04 to -0.01, *p* < 0.001 to *p* < .05).

### Term/Incomplete Pregnancies as Categorical Variables

Results were consistent when using term/incomplete pregnancies as categorical variables, with the strongest association between term pregnancy and DSST scores in women who reported 5 or more term pregnancies compared to women with no reported term pregnancies (β = -0.46 to -0.10, *p* = < 0.001 to 0.3), no other categorical groups were significantly different from the reference group for DSST scores. We also found a difference in AF scores between the reference group and participants that reported 1-2 term pregnancies and 3-4 term pregnancies but not those that reported 5 or more term pregnancies. No significant associations were found for AF scores and incomplete pregnancies in any of our categorical groups. In line with the analysis using continuous term and incomplete pregnancy variables, the relationship between CERAD-WL and term/incomplete pregnancies were not significant across any of the categorical groups. Interestingly, the relationship between both term pregnancies and incomplete pregnancies and CERAD-DR scores was no longer significant when examining term and incomplete pregnancies as categorical variables. Results for models using term and incomplete pregnancy as categorical variables are found in Supplemental Table 1.

### Formal Mediation Analysis

In the formal mediation analysis, the association between term pregnancies and DSST, AF, and CERAD-DR scores were significantly mediated by PIR but not education level. The indirect effect of term pregnancies on DSST mediated through PIR was statistically significant (β = -0.02, 95% CI = -0.03 to -0.02), which can be interpreted as 28% of the effect of term pregnancies on DSST mediated through PIR. Additionally, we found that the proportion mediated in the relationships between term pregnancies with AF (β = -0.01, 95% CI = -0.02 to - 0.01) and CERAD-DR (β = -0.01, 95% CI = -0.02 to -0.01) to be 34% and 26%, respectively. The indirect effects of term pregnancies on DDST (β = -0.02, 95% CI = -0.06 to 0.02), AF (β = - 0.01, 95% CI = -0.04 to 0.00), and CERAD-DL (β = -0.02, 95% CI = -0.04 to 0.01) mediated through education level was not statistically significant. Full results for the mediating effect of PIR and education level on the relationship between term pregnancy and all cognitive tests can be found in Tables 3 and 4.

## DISCUSSION

We found that postmenopausal women who had more term pregnancies, showed significantly worse cognitive performance on the DSST and CERAD-WL; however, this association was attenuated when adjusting for PIR and education level. We found positive relationships between incomplete pregnancies and CERAD-DR scores. When using term and incomplete pregnancies as categorical variables, we found a significant association between term pregnancy and DSST scores in women who reported 5 or more term pregnancies compared to women who reported no term pregnancies. We found strong associations between AF scores and term pregnancies in participants that reported 1-2 term pregnancies and 3-4 term pregnancies but not in those that reported 5 or more term pregnancies. In our formal mediation analysis we found that DSST, AF, and CERAD-DR scores were mediated by PIR but not education level. No other associations between pregnancy history and cognitive function were observed.

Our results differed with the findings from a prior study from Tsai et al^4^, who, using NHANES data from different waves corresponding to years 1999-2002, with similar demographic characteristics and covariates, found that women who had 5 or more pregnancies demonstrated lower DSST scores even after adjusting for relevant covariates.^4^ There are several explanations for the differences between our findings and this prior study. First, Tsai et al^4^ pregnancy groupings were as follows: 0-1, 2-3, and 5 or more. In our approach, we grouped number of pregnancies in a unique way; our pregnancy groups separate out individuals who reported no term pregnancies (0 term pregnancies) or no incomplete pregnancies (0 incomplete pregnancies) from those who reported term pregnancies (1-2, 3-4, 5 or more) or incomplete pregnancies (1-2, 3 or more). Moreover, we separately examined number of term pregnancies and number of incomplete pregnancies in order to determine whether these different types of pregnancies relate differently to cognitive function. Second, the analysis done by Tsai et al^4^ only used pregnancy as a categorical variable, our models were run using the aforementioned categorical pregnancy variables as well as a continuous version for each of these variables. Finally, we utilized a more complex education variable that included the following levels: Less than 9th grade, 9-11th grade, High School/GED, some college, and college graduate or above; this is a distinct difference from the Tsai et al^4^ variable that included only two levels, more or less than 12 years of education. Given the differences in variables and analysis techniques between Tsai et al^4^ and our approach we expected, to some extent, to find differences between our results.

We observed a slightly positive relationship between incomplete pregnancy and CERAD-DR scores. Previous studies have reported a positive association of incomplete pregnancies with cognition; however, both of these studies relied upon one cognitive measure (MMSE and CDR) to classify cognition. These measures provide an overall idea of cognitive status but little specificity as to which specific cognitive abilities are altered.^5,18^ Given that significant associations were only found between incomplete pregnancies and one cognitive test in our study, indicates further research is needed to understand the relationship between cognitive function and history of incomplete pregnancies.

Our results support previous research that has indicated socioeconomic indicators like education and income level play an important role in cognitive performance in later life.^39–41^ We found that PIR mediated the relationship of term pregnancy history and multiple domains of cognitive performance, but not education level. We suspected the reason that education level did not significantly mediate the relationship between term pregnancy and cognition was due to the temporality of our education variable, as we assume the highest level of education was most likely achieved before these women were pregnant. Our results indicate a necessity to take SES factors into consideration when conducting these types of analysis and also highlight the importance of programs/interventions aimed at reducing the effects of poverty on families.

There are several limitations of the present study that should be acknowledged. First, many of our variables (e.g., age of first period, age of menopause, number of pregnancies) were collected by self-report when women were postmenopausal and thus may be vulnerable to recall bias. Regarding the calculation of term pregnancies and incomplete pregnancies, it is important to note that these variables are imperfect measures of pregnancy and extremely complicated variables to define. For example, due to limitations in the NHNANES protocol, it was not possible to distinguish between the many unique pregnancy situations that arise such as twins, children born prematurely, abortions, spontaneous incomplete pregnancies, and recurrent incomplete pregnancies. Furthermore, the time of miscarriage or details regarding miscarriage were not available in this dataset. Without this information, it makes it difficult to draw conclusions on the implications of incomplete pregnancy as we are unsure in what trimester the pregnancy ended. This lack of detail makes it hard to understand the biological underpinnings of the relationship between pregnancy and cognition. Finally, cognitive function was only measured by three cognitive tasks; thus, we were unable to assess a comprehensive number of cognitive domains.

Future studies would benefit from including questions about pregnancy complications during the reproductive history interview. Women who report preeclampsia and gestational diabetes have shown higher rates of cardiovascular diseases after pregnancy and throughout their life. This is important as these conditions could be a potential risk factor for cognitive decline later in life.^42^ Additionally, future research should consider the role of APOE in the association of pregnancy with cognition. Corbo et al^43^ found that pregnancy was associated with lower AD onset in participants carrying ApoE3 genotype but not ApoE4 genotype carriers, suggesting that pregnancy interacts with specific genotypes in the context of cognitive aging.^43^

## CONCLUSIONS

Women with a greater number of term pregnancies were found to have worse cognitive performance, whereas a greater number of incomplete pregnancies was associated with better cognitive performance. Our results were partially mediated by the effects of PIR. These findings suggest that further research is needed that takes into consideration variables that account for both social and biological factors to fully understand the complexity of the relationship between pregnancy and cognition in women.

## Data Availability

Data used in this study and corresponding protocols on data collection is freely available on the National Health and Nutrition Survey website.

https://www.cdc.gov/nchs/nhanes/index.htm

## Table/Figure Legends

Table 1: Demographics of Study Sample

Table 2: Regression Models for Continuous Pregnancy Variables

Table 3: Mediation Analysis of PIR for Number of Pregnancies by Cognitive Test

Table 4: Mediation Analysis of Education Level for Number of Pregnancies by Cognitive Test

Supplemental Table 1: Regression Models for Categorical Pregnancy Variables

**Nonstandard Characters:**

β = Beta Level

## Notes

**Financial Support:** NIH/NIA AG063843 awarded to Dr. Panizzon NIH 1R01AG066088-01and CDPH-19-10613 awarded to Dr. Banks NIH R01AG074221 and CDPH-19-10613 awarded to Dr. Sundermann

**Conflict of Interest/Financial Disclosure:** None reported

### Competing Interest Statement

The authors have declared no competing interest.

### Clinical Trial

Study was conducted using secondary data analysis.

### Funding Statement

NIH/NIA AG063843 awarded to Dr. Panizzon
NIH 1R01AG066088-01and CDPH-19-10613 awarded to Dr. Banks
NIH R01AG074221 and CDPH-19-10613 awarded to Dr. Sundermann

### Author Declarations

This study was done through secondary data analysis, proper IRB procedures were completed by NHANES study during the data collection process.

