## Supplementary material for "The Mediating Role of SES on the Relationship between Pregnancy History and Later Life Cognition": Tables

**Table 1: Demographics of Study Sample**

| Number of Term Pregnancies | Overall, N = 1,016 | 0, N = 116 | 1-2, N = 420 | 3-4, N = 339 | 5 or more, N = 141 | p-value[a] |
|---|---|---|---|---|---|---|
| Age, Mean (SD) | 67.3 (5.4) | 67.1 (5.4) | 66.4 (5.0) | 67.7 (5.5) | 69.3 (5.6) | **<0.001** |
| Race, n (%) | | | | | | **<0.001** |
| Hispanic | 206 (20%) | 16 (14%) | 67 (16%) | 77 (23%) | 46 (33%) | |
| White | 466 (46%) | 57 (49%) | 212 (50%) | 151 (45%) | 46 (33%) | |
| Black | 240 (24%) | 23 (20%) | 90 (21%) | 86 (25%) | 41 (29%) | |
| Asian | 90 (8.9%) | 16 (14%) | 47 (11%) | 22 (6.5%) | 5 (3.5%) | |
| Multi-Cultural | 14 (1.4%) | 4 (3.4%) | 4 (1.0%) | 3 (0.9%) | 3 (2.1%) | |
| Education Level, n (%) | | | | | | **<0.001** |
| High school graduate/GED | 249 (25%) | 22 (19%) | 101 (24%) | 92 (27%) | 34 (24%) | |
| Less than 9th grade | 90 (8.9%) | 7 (6.0%) | 18 (4.3%) | 28 (8.3%) | 37 (26%) | |
| 9-11th grade | 138 (14%) | 6 (5.2%) | 37 (8.8%) | 62 (18%) | 33 (23%) | |
| Some College | 323 (32%) | 34 (29%) | 154 (37%) | 107 (32%) | 28 (20%) | |
| College graduate or above | 216 (21%) | 47 (41%) | 110 (26%) | 50 (15%) | 9 (6.4%) | |
| PIR, Mean (SD) | 2.58 (1.62) | 2.78 (1.69) | 2.95 (1.69) | 2.42 (1.50) | 1.68 (1.21) | **<0.001** |
| Reproductive Span, Mean (SD) | 33 (8) | 34 (8) | 34 (8) | 33 (8) | 33 (7) | 0.2 |
| BMI, Mean (SD) | 30 (7) | 29 (8) | 30 (8) | 30 (7) | 31 (7) | **0.048** |
| Smoked at least 100 cigarettes in life?, | 400 (39%) | 50 (43%) | 169 (40%) | 125 (37%) | 56 (40%) | 0.6 |
| PHQ (Depression Screener), Mean (SD) | 3.9 (4.9) | 3.4 (4.6) | 3.7 (4.7) | 4.0 (4.9) | 4.9 (5.6) | 0.2 |
| Medical Conditions, Mean (SD) | 1.34 (1.06) | 1.29 (1.01) | 1.27 (1.03) | 1.33 (1.07) | 1.61 (1.14) | **0.017** |
| Incomplete Pregnancies, n (%) | | | | | | 0.081 |
| 0 | 645 (63%) | 83 (72%) | 264 (63%) | 215 (63%) | 83 (59%) | |
| 1-2 | 322 (32%) | 27 (23%) | 135 (32%) | 114 (34%) | 46 (33%) | |
| 3 or more | 49 (4.8%) | 6 (5.2%) | 21 (5.0%) | 10 (2.9%) | 12 (8.5%) | |

[a]Kruskal-Wallis rank sum test; Pearson's Chi-squared test

Note: PIR = Poverty Index Ratio; Reproductive Span = Age Menopause--Age of Menarche; PHQ = Patient Health Questionnaire (Depression Screener); Medical Conditions = Diabetes + Heart Disease + Thyroid Disease + History of Stroke + Sleep Disorders + Hypertension. Values: 0=No, 1=Yes, 0.5=Borderline (diabetes only), the higher an individual's score the higher number of the aforementioned conditions participant self-reported.

**Table 2: Linear Regression Continuous Pregnancy Variables**

| | | Model 1 | | | Model 2 | | | Model 3 | | | Model 4 | | |
|---|---|---|---|---|---|---|---|---|---|---|---|---|---|
| Cognitive Test | Type of Pregnancy | Beta[a] | SE[b] | 95% CI[b] | Beta[a] | SE[b] | 95% CI[b] | Beta[a] | SE[b] | 95% CI[b] | Beta[a] | SE[b] | 95% CI[b] |
| DSST | Term | -0.09*** | 0.014 | -0.12, -0.06 | -0.03* | 0.014 | -0.06, -0.01 | -0.07*** | 0.014 | -0.09, -0.04 | -0.03 | 0.013 | -0.05, 0.00 |
| | Incomplete | 0.01 | 0.028 | -0.05, 0.06 | 0 | 0.025 | -0.05, 0.05 | 0.01 | 0.027 | -0.04, 0.06 | 0.01 | 0.025 | -0.04, 0.05 |
| | $R^2$ | 0.329 | | | 0.455 | | | 0.386 | | | 0.474 | | |
| AF | Term | -0.03* | 0.016 | -0.07, 0.00 | 0 | 0.016 | -0.03, 0.03 | -0.02 | 0.016 | -0.05, 0.01 | 0 | 0.016 | -0.03, 0.03 |
| | Incomplete | 0.06* | 0.031 | 0.00, 0.12 | 0.06 | 0.03 | 0.00, 0.11 | 0.06* | 0.03 | 0.00, 0.12 | 0.06 | 0.03 | 0.00, 0.12 |
| | $R^2$ | 0.189 | | | 0.24 | | | 0.203 | | | 0.241 | | |
| CERAD-WL | Term | -0.02 | 0.016 | -0.05, 0.01 | 0.01 | 0.017 | -0.02, 0.04 | -0.01 | 0.016 | -0.04, 0.02 | 0.01 | 0.017 | -0.02, 0.05 |
| | Incomplete | 0.05 | 0.031 | -0.01, 0.11 | 0.05 | 0.031 | -0.01, 0.11 | 0.05 | 0.031 | -0.01, 0.11 | 0.05 | 0.03 | -0.01, 0.11 |
| | $R^2$ | 0.126 | | | 0.176 | | | 0.141 | | | 0.179 | | |
| CERAD-DR | Term | -0.04** | 0.017 | -0.08, -0.01 | -0.02 | 0.017 | -0.05, 0.02 | -0.03* | 0.017 | -0.07, 0.00 | -0.01 | 0.017 | -0.05, 0.02 |
| | Incomplete | 0.07* | 0.032 | 0.01, 0.13 | 0.07* | 0.031 | 0.00, 0.13 | 0.07* | 0.032 | 0.01, 0.13 | 0.07* | 0.031 | 0.01, 0.13 |
| | $R^2$ | 0.119 | | | 0.149 | | | 0.132 | | | 0.153 | | |

[a]*p<0.05; **p<0.01; ***p<0.001

[b]SE = Standard Error, CI = Confidence Interval

**Model 1/Base Model**: Age at Screening, Race, Length of Reproductive Span, BMI, Smoking Status, Medical Conditions Score, and PHQ (depression screener); **Model 2:** Model 1 + Education Level; **Model 3:** Model 1 + Federal Income-to-Poverty Ratio (PIR); **Model 4:** Model 1 + Education Level and PIR

**DSST:** Digit Symbol Substitution Test, **AF:** Animal Fluency, **CERAD-WL:** CERAD Word Learning, **CERAD-DR:** CERAD Delayed Recall

**Table 3:** Mediation Analysis of PIR for Number of Pregnancies by Cognitive Test

| | Estimate | 95% CI Lower | 95% CI Upper | Proportion Mediated (95%CI) |
|---|---|---|---|---|
| **DSST** | | | | 0.28 (0.17-0.42) |
| Average Indirect Effects | -0.02 | -0.03 | -0.02 | |
| Average Direct Effects | -0.07 | -0.09 | -0.04 | |
| Total Effect | -0.09 | -0.12 | -0.06 | |
| **AF** | | | | 0.34 (0.03-1.84) |
| Average Indirect Effects | -0.01 | -0.02 | -0.01 | |
| Average Direct Effects | -0.02 | -0.06 | 0.01 | |
| Total Effect | -0.03 | -0.07 | 0.00 | |
| **CERAD-WL** | | | | 0.56 (-4.60-5.96) |
| Average Indirect Effects | -0.01 | -0.02 | -0.01 | |
| Average Direct Effects | -0.01 | -0.05 | 0.03 | |
| Total Effect | -0.02 | -0.06 | 0.02 | |
| **CERAD-DR** | | | | 0.26 (0.10-1.48) |
| Average Indirect Effects | -0.01 | -0.02 | -0.01 | |
| Average Direct Effects | -0.03 | -0.07 | 0.01 | |
| Total Effect | -0.04 | -0.08 | -0.01 | |

Note: **DSST:** Digit Symbol Substitution Test, **AF:** Animal Fluency, **CERAD-WL:** CERAD Word Learning, **CERAD-DR:** CERAD Delayed Recall

**Table 4:** Mediation Analysis of Education Level for Number of Pregnancies by Cognitive Test

| | Estimate | 95% CI Lower | 95% CI Upper | Proportion Mediated (95%CI) |
|---|---|---|---|---|
| **DSST** | | | | 0.42 (-0.50-0.94) |
| Average Indirect Effects | -0.02 | -0.06 | 0.02 | |
| Average Direct Effects | -0.03 | -0.06 | 0.00 | |
| Total Effect | -0.06 | -0.10 | -0.01 | |
| **AF** | | | | 1.14 (-7.64-8.13) |
| Average Indirect Effects | -0.01 | -0.04 | 0.00 | |
| Average Direct Effects | 0.00 | -0.03 | 0.04 | |
| Total Effect | -0.01 | -0.06 | 0.02 | |
| **CERAD-WL** | | | | 3.10 (-7.89-12.36) |
| Average Indirect Effects | -0.02 | -0.04 | 0.01 | |
| Average Direct Effects | 0.01 | -0.03 | 0.05 | |
| Total Effect | -0.01 | -0.05 | 0.04 | |
| Prop. Mediated | 2.05 | -7.88 | 15.39 | |
| **CERAD-DR** | | | | 0.14 (-1.94-2.85) |
| Average Indirect Effects | 0.00 | -0.03 | 0.00 | |
| Average Direct Effects | -0.02 | -0.06 | 0.02 | |
| Total Effect | -0.02 | -0.07 | 0.01 | |

Note: **DSST:** Digit Symbol Substitution Test, **AF:** Animal Fluency, **CERAD-WL:** CERAD Word Learning, CERAD-DR: CERAD Delayed Recall

**Supplemental Table 1: Linear Regression Categorical Pregnancy Variables**

| | | Model 1 | | | Model 2 | | | Model 3 | | | Model 4 | | |
|---|---|---|---|---|---|---|---|---|---|---|---|---|---|
| Cognitive Exercise | Type of Pregnancy | Betaa[a] | SE[b] | 95% CI[b] | Beta[a] | SE[b] | 95% CI[b] | Beta[a] | SE[b] | 95% CI[b] | Beta[a] | SE[b] | 95% CI[b] |
| **DSST** | **Term** | | | | | | | | | | | | |
| | 0 | — | — | — | — | — | — | — | — | — | — | — | — |
| | 1-2 | -0.05 | 0.087 | -0.22, 0.12 | -0.02 | 0.078 | -0.17, 0.14 | -0.08 | 0.083 | -0.24, 0.08 | -0.05 | 0.077 | -0.20, 0.10 |
| | 3-4 | -0.16 | 0.09 | -0.33, 0.02 | -0.02 | 0.082 | -0.18, 0.14 | -0.12 | 0.086 | -0.28, 0.05 | -0.03 | 0.081 | -0.19, 0.13 |
| | 5 or more | -0.46*** | 0.106 | -0.67, -0.26 | -0.12 | 0.098 | -0.31, 0.07 | -0.34** | 0.102 | -0.54, -0.14 | -0.1 | 0.097 | -0.29, 0.09 |
| | **Incomplete** | | | | | | | | | | | | |
| | 0 | — | — | — | — | — | — | — | — | — | — | — | — |
| | 1-2 | 0.05 | 0.057 | -0.07, 0.16 | 0.02 | 0.052 | -0.08, 0.12 | 0.05 | 0.054 | -0.06, 0.15 | 0.02 | 0.051 | -0.08, 0.12 |
| | 3 or more | -0.09 | 0.126 | -0.34, 0.15 | -0.06 | 0.113 | -0.28, 0.16 | -0.05 | 0.12 | -0.29, 0.18 | -0.04 | 0.112 | -0.26, 0.18 |
| | $R^2$ | 0.323 | | | 0.454 | | | 0.382 | | | 0.472 | | |
| **AF** | **Term** | | | | | | | | | | | | |
| | 0 | — | — | — | — | — | — | — | — | — | — | — | — |
| | 1-2 | 0.16 | 0.095 | -0.02, 0.35 | 0.21* | 0.092 | 0.03, 0.39 | -0.02 | 0.096 | -0.21, 0.17 | 0.21* | 0.093 | 0.03, 0.39 |

| | | | | | | | | | | | | | |
|---|---|---|---|---|---|---|---|---|---|---|---|---|---|
| | 3-4 | 0.07 | 0.098 | -0.12, 0.26 | 0.19* | 0.097 | 0.00, 0.38 | 0.03 | 0.1 | -0.17, 0.22 | 0.19* | 0.097 | 0.00, 0.38 |
| | 5 or more | -0.11 | 0.116 | -0.33, 0.12 | 0.12 | 0.116 | -0.11, 0.34 | 0.01 | 0.119 | -0.22, 0.24 | 0.12 | 0.116 | -0.11, 0.35 |
| | **Incomplete** | | | | | | | | | | | | |
| | 0 | — | — | — | — | — | — | — | — | — | — | — | — |
| | 1-2 | 0.12 | 0.062 | 0.00, 0.24 | 0.09 | 0.061 | -0.03, 0.21 | 0.04 | 0.063 | -0.08, 0.17 | 0.09 | 0.061 | -0.02, 0.21 |
| | 3 or more | 0.14 | 0.137 | -0.13, 0.41 | 0.15 | 0.133 | -0.11, 0.41 | 0.2 | 0.14 | -0.08, 0.47 | 0.15 | 0.133 | -0.11, 0.42 |
| | $R^2$ | 0.194 | | | 0.244 | | | 0.14 | | | 0.245 | | |
| **CERAD-WL** | **Term** | | | | | | | | | | | | |
| | 0 | — | — | — | — | — | — | — | — | — | — | — | — |
| | 1-2 | 0 | 0.097 | -0.19, 0.19 | 0.03 | 0.095 | -0.16, 0.21 | -0.02 | 0.096 | -0.21, 0.17 | 0.01 | 0.095 | -0.17, 0.20 |
| | 3-4 | 0.01 | 0.1 | -0.19, 0.20 | 0.09 | 0.099 | -0.11, 0.28 | 0.03 | 0.1 | -0.17, 0.22 | 0.08 | 0.099 | -0.11, 0.28 |
| | 5 or more | -0.05 | 0.119 | -0.29, 0.18 | 0.15 | 0.119 | -0.08, 0.39 | 0.01 | 0.119 | -0.22, 0.24 | 0.16 | 0.119 | -0.07, 0.40 |
| | **Incomplete** | | | | | | | | | | | | |
| | 0 | — | — | — | — | — | — | — | — | — | — | — | — |
| | 1-2 | 0.04 | 0.064 | -0.08, 0.17 | 0.03 | 0.062 | -0.09, 0.16 | 0.04 | 0.063 | -0.08, 0.17 | 0.04 | 0.062 | -0.09, 0.16 |

| | | | | | | | | | | | | | |
|---|---|---|---|---|---|---|---|---|---|---|---|---|---|
| | 3 or more | 0.18 | 0.141 | -0.10, 0.45 | 0.2 | 0.137 | -0.07, 0.47 | 0.2 | 0.14 | -0.08, 0.47 | 0.21 | 0.137 | -0.06, 0.48 |
| | $R^2$ | 0.124 | | | 0.177 | | | 0.14 | | | 0.181 | | |
| **CERAD-DR** | **Term** | | | | | | | | | | | | |
| | 0 | — | — | — | — | — | — | — | — | — | — | — | — |
| | 1-2 | -0.02 | 0.099 | -0.22, 0.17 | 0 | 0.098 | -0.19, 0.20 | -0.04 | 0.098 | -0.23, 0.16 | -0.01 | 0.098 | -0.20, 0.18 |
| | 3-4 | -0.1 | 0.102 | -0.30, 0.10 | -0.03 | 0.102 | -0.23, 0.17 | -0.08 | 0.102 | -0.28, 0.12 | -0.03 | 0.102 | -0.23, 0.17 |
| | 5 or more | -0.21 | 0.121 | -0.45, 0.03 | -0.04 | 0.123 | -0.28, 0.20 | -0.15 | 0.121 | -0.38, 0.09 | -0.03 | 0.123 | -0.27, 0.21 |
| | **Incomplete** | | | | | | | | | | | | |
| | 0 | — | — | — | — | — | — | — | — | — | — | — | — |
| | 1-2 | 0.1 | 0.065 | -0.03, 0.23 | 0.09 | 0.064 | -0.04, 0.21 | 0.1 | 0.065 | -0.03, 0.23 | 0.09 | 0.064 | -0.04, 0.21 |
| | 3 or more | 0.18 | 0.144 | -0.10, 0.47 | 0.2 | 0.141 | -0.08, 0.48 | 0.21 | 0.143 | -0.07, 0.49 | 0.21 | 0.141 | -0.07, 0.49 |
| | $R^2$ | 0.115 | | | 0.148 | | | 0.129 | | | 0.152 | | |

[a]*p<0.05; **p<0.01; ***p<0.001

[b]SE = Standard Error, CI = Confidence Interval

Note: 0 Term or incomplete pregnancies are the reference groups; **DSST:** Digit Symbol Substitution Test, **AF:** Animal Fluency, **CERAD-WL:** CERAD Word Learning, **CERAD-DR:** CERAD Delayed Recall
